# Social determinants of health and epigenetic clocks: Meta-analysis of 140 studies

**DOI:** 10.1101/2025.05.08.25327207

**Authors:** Y.E. Willems, A. D. Rezaki, M. Aikins, A. Bahl, Q. Wu, D.W. Belsky, L. Raffington

## Abstract

Social determinants of health are social factors that affect health and survival. Two of the most powerful social determinants are socioeconomic status (SES) and race/ethnicity; people with lower SES or marginalized race/ethnicity tend to experience earlier onset of aging-related diseases and have shorter lifespans. DNA methylation (DNAm) measures of biological aging, often referred to as “epigenetic clocks”, are increasingly used to study the social determination of health. However, there are several generations of epigenetic clocks and it remains unclear which are most sensitive to social factors affecting health. Moreover, there is uncertainty about how technical factors, such as the tissue from which DNA is derived or the technology used to measure DNA methylation may affect associations of social determinants with epigenetic clocks. We conducted a pre-registered multi-level meta-analysis of 140 studies, including N = 65,919 participants, encompassing 1,065 effect sizes for associations of SES and racial/ethnic identity with three generations of epigenetic clocks. We found that associations were weakest for the first generation of epigenetic clocks developed to predict age differences between people. Associations were stronger for the second generation of epigenetic clocks developed to predict mortality and health risks. The strongest associations were observed for a third generation of epigenetic clocks, sometimes referred to as “epigenetic speedometers”, developed to predict the pace of aging. In studies of children, only the speedometers showed significant associations with SES. Effects of sex and technical factors were minimal and there was no evidence of publication bias.

## Introduction

Individuals with lower socioeconomic status (SES) or who have marginalized racial and ethnic identities often experience an earlier onset of chronic diseases and a shorter lifespan compared to their more advantaged counterparts ^1,2^. One hypothesis advanced to explain these disparities is that social disadvantage accelerates processes of biological aging ^3–7^. Biological aging is the progressive loss of integrity and resilience capacity in cells, tissues, and organs with the passage of time ^5,8^. Biological aging is thought to arise from an accumulation of molecular damage resulting in “hallmark” alternations to the structure and function of our cells that ultimately cause many different chronic diseases and underpin the general decline in functional capacity that occurs with aging ^9–11^. While there is no gold standard measurement of biological aging ^8,12^, a family of machine-learning-derived DNA methylation (DNAm) algorithms known as “epigenetic clocks” are among the most widely studied and best-validated predictors of healthy lifespan across a range of human populations^13,14^. They are emerging as an important tool in social determinants of health research, where researchers are using them to track the biological embedding of health risks, especially among individuals who have not yet developed aging-related chronic health conditions.

Epigenetic clocks are developed by modeling proxy measures of biological aging from whole-genome DNA methylation patterns. The result is a predictive algorithm that, when applied to DNA methylation data from a new sample, returns a numeric value representing the progress or pace of biological aging. Within this general approach, there are now multiple “generations” of clocks defined by the proxy measure of aging used to develop the algorithm ^3,15^. The first generation of epigenetic clocks was developed by modeling differences between individuals in their chronological age *(i.e.* time since birth) ^16,17^. First-generation epigenetic clocks estimate biological age as the age at which a person’s DNA methylation pattern matches the norm in a reference population. While first-generation clocks accurately estimate chronological age, their ability to predict health outcomes appears to be more limited ^18^. To overcome this limitation, the second generation of epigenetic clocks was developed by modeling differences between individuals in their risk of death and translating these risks into age-equivalent values. Second-generation epigenetic clocks estimate biological age as the age at which an individual’s predicted mortality and health risk match the norm in a reference population ^19,20^. A third generation of epigenetic clocks was developed by modeling differences between individuals in the rate at which their bodies were deteriorating over time. Rather than estimating a biological age, these “epigenetic speedometers” estimate a person’s Pace of Aging, or the rate at which biological age is increasing relative to the norm in a reference population ^21,22^.

A growing body of research indicates that epigenetic clocks are sensitive to socioeconomic, racial, and ethnic disparities ^23–27^. However, there are a large number of clocks and they do not all yield the same information. In fact, correlations across clocks developed using different generations of methods tend to be modest ^25,28^. Key knowledge gaps include which clocks are most sensitive to social determinants of health, when during the life course socioeconomic exposures significantly impact biological aging, whether there are potential sex differences in these relations, and how to determine the most critical technical considerations for analysis. In this context, researchers need guidance on selecting clocks that rigorously test hypotheses related to social determinants of health and identifying the life stages where these associations are strongest. Addressing these questions will enable researchers to incorporate the most suitable clocks into quasi-experimental and randomized controlled studies exploring potential anti-aging interventions.

We conducted a pre-registered, multi-level meta-analysis synthesizing findings on the associations of SES and race/ethnicity with three generations of epigenetic-clock measures of biological aging (https://osf.io/u5z82). *First*, we examined SES associations with epigenetic clocks. *Second*, we investigated whether these associations varied depending on when in life (childhood vs. adulthood) SES and DNAm were measured. *Third*, we examined race/ethnicity associations with epigenetic clocks. *Fourth*, we explored potential sex differences in associations with epigenetic clocks. *Fifth*, we evaluated whether technical factors, including tissue type and array platform, impacted these associations, and whether there was evidence of publication bias.

## Results

### Descriptive Results

Figure 1 presents the workflow of our analysis, which is described in the Methods section. Our final meta-analytic dataset consisted of 140 studies including N = 65,919 unique participants and a total of 1,065 effect sizes derived from 79 unique cohorts drawn from 23 different countries (Supplemental Figure 1). Figure 2 is a PRISMA flowchart depicting the process of selecting studies. The majority of cohorts (52%) were from the USA, followed by the UK (9%). More than half of the cohorts (57%) were based on predominantly (>60%) White samples, and the majority (77%) had a mixed distribution of males and females. Sample sizes across all cohorts ranged from n=33 to n=16,245. The mean age was 49.9 years, ranging from 0 - 86 years. Figure 3 shows a heatmap of effect sizes across socioeconomic and race/ethnicity measures with the darker shades corresponding to larger numbers of effect-size estimates included in the meta-analysis. The full meta-analytic dataset and analytic scripts are available as supplements. Deviations from pre-registration are described in Supplemental Table 1.

**Figure 1.**
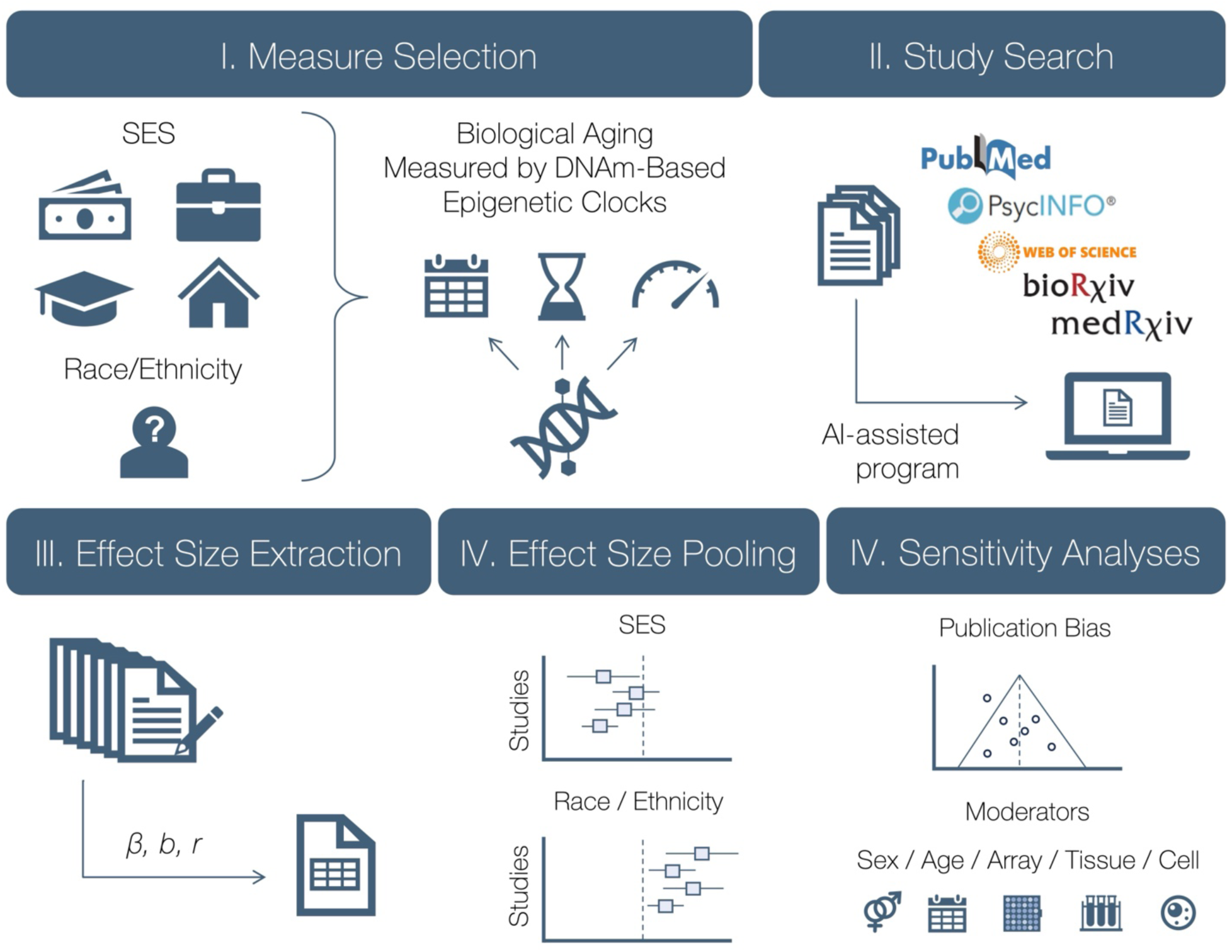
Methodological workflow of the present meta-analysis. *Panel I* visualizes measures of interest, including socioeconomic status (SES), race and ethnicity, and epigenetic-clock measures of biological aging. *Panel II* depicts the study search process across PubMed, PsycINFO, Web of Science, bioRxiv and medRxiv databases. The results were screened at the title and abstract level using an AI-assisted program and then screened at the full-text level by two researchers. *Panel III* indicates that effect sizes, along with information on the study and cohort characteristics, were extracted and harmonized from the selected articles by three authors. *Panel IV* shows effect sizes were converted to a common metric and pooled. The associations of SES and self-reported race/ethnicity with epigenetic-clock-measured biological aging were pooled and meta-analyzed. *Panel V* details moderator analyses for pre-registered technical and theoretical moderators as well as sensitivity analyses, including probing evidence for publication bias.

**Figure 2.**
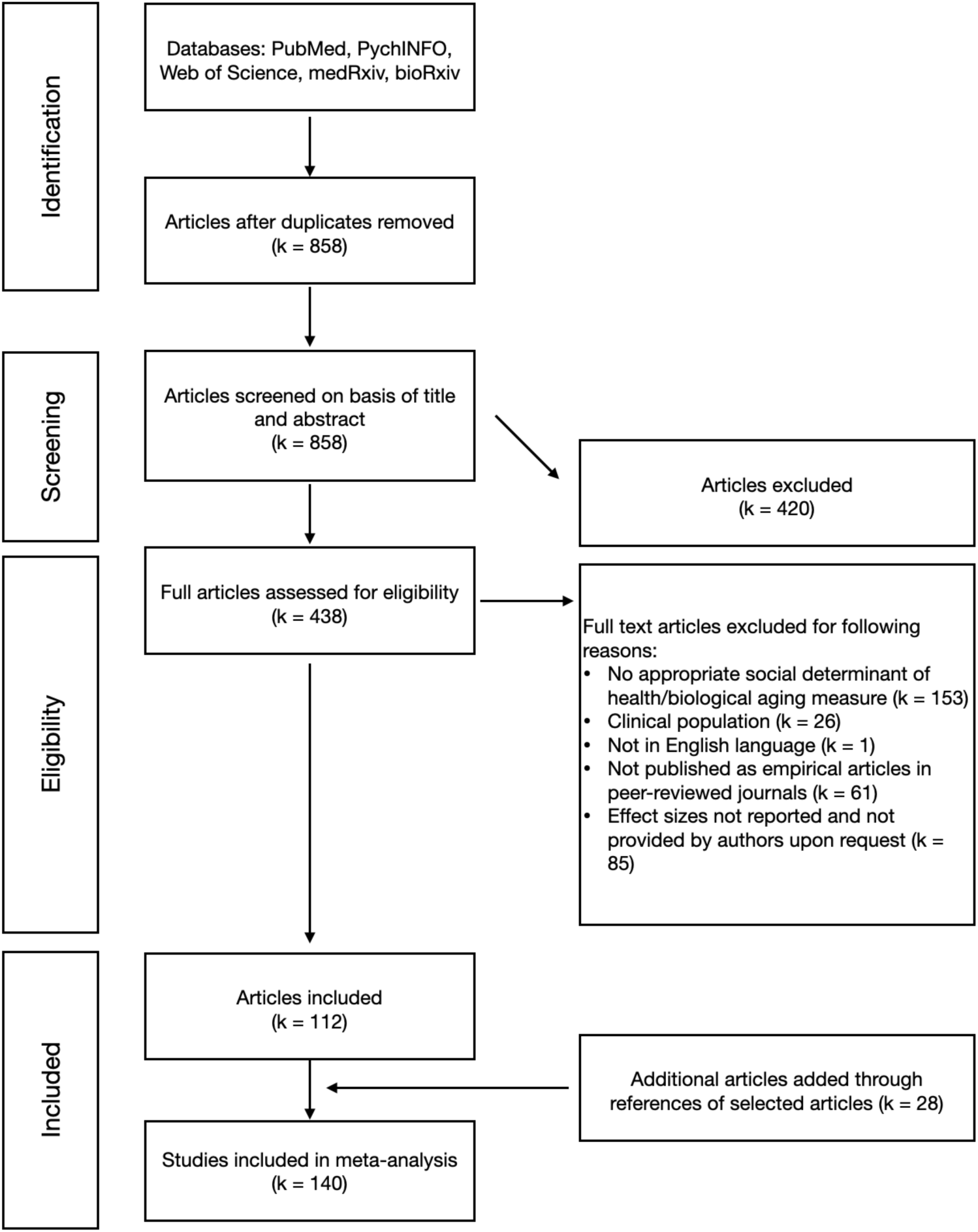
PRISMA flow diagram. The search conducted from 2013 to February 2024.

**Figure 3.**
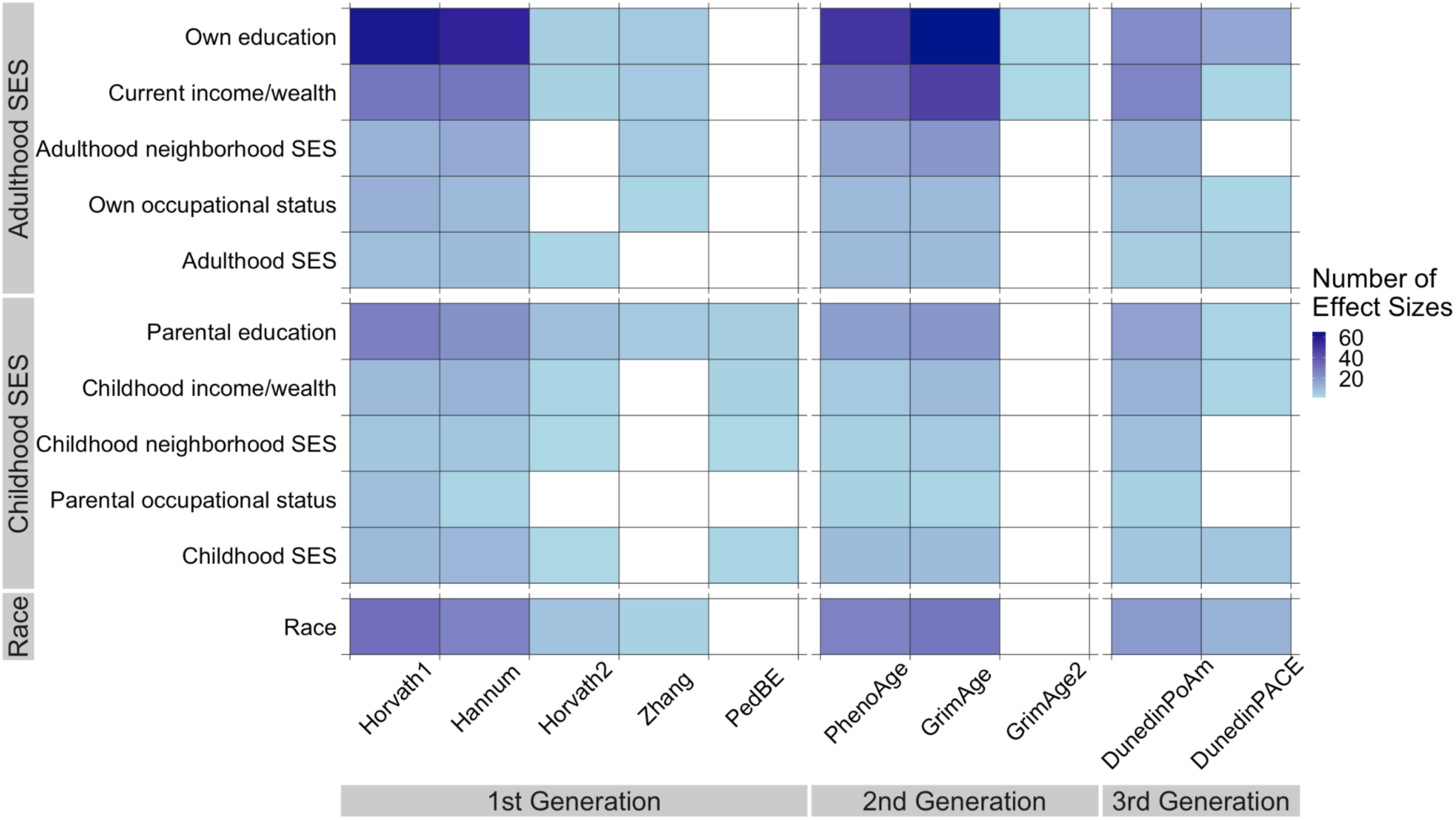
Heatmap showing the number of effect sizes for epigenetic-clock-measured biological aging (x-axis) and social determinants of health (y-axis). Number of effect sizes per category ranges from 60 (dark blue) to 1 (light blue). White squares indicate no effect sizes.

### (1) Associations of SES with biological aging by epigenetic clock generation and measure

We first examined whether epigenetic-clock-measured biological aging was associated with SES, considering the influence of clock generation (*i.e.,* first-generation clocks developed to predict chronological age; second-generation clocks developed to predict mortality and health risk on blood chemistry and age data; third-generation clocks developed to predict the pace of aging) and clock measure (*e.g.,* Horvath, GrimAge, DunedinPACE). For this analysis, we conducted a statistical test of effect-measure-modification (moderation) of SES-clock associations by clock generation or measure. Effect sizes included in this analysis were derived from 23 countries including individuals from birth to old age and various measures of SES (*e.g.,* own education, parental income, composite neighborhood socioeconomic status, see Table 1). Effect sizes are reported as Pearson’s r with 95% confidence intervals.

**Table 1.** Overview of key concepts of interest for Epigenetic-clock-measured biological aging and social determinants of health.

| Epigenetic-clock-measured biological aging | Social determinants of health |
| --- | --- |
| <u>1st generation measures:</u><br>- Horvath pan-tissue clock<sup>17</sup><br>- Hannum clock<sup>16</sup><br>- Horvath Skin and Blood clock<sup>74</sup><br>- Zhang Clock<sup>75</sup><br>- PedBE Clock<sup>76</sup> | <u>Socioeconomic status in adulthood:</u><br>- income/wealth<br>- education<br>- occupational status<br>- composite measures that combine two or more of these variables into one SES measure<br>- neighborhood SES |
| <u>2nd generation measures:</u><br>- PhenoAge<sup>19</sup><br>- GrimAge<sup>20,77</sup> | <u>Socioeconomic status in childhood:</u><br>- parental income/wealth<br>- parental education<br>- parental occupation<br>- composite measures that combine two or more of these variables into one SES measure<br>- neighborhood SES |
| <u>3rd generation measures:</u><br>- DunedinPoAm<sup>21</sup><br>- DunedinPACE<sup>22</sup> | <u>Racial/ethnicity</u><br>- Self-reported racial and ethnic identity |

We found that individuals of lower SES tended to exhibit accelerated and faster epigenetic-clock-measured biological aging compared to individuals of higher SES. The magnitude of this association between SES and biological aging was moderated by clock generation (*F*(2) = 178.10, *p* < .001) and clock measure (*F*(8) = 244.88, *p* < .001, Supplemental Table 1A). It was largest for third-generation measures (*r* = −0.13 [−0.15, −0.11]) followed by second-generation measures (*r* = −0.11 [−0.12, −0.09]), both of which were larger than first-generation measures (*r* = −0.03 [−0.04, −0.01]; see Figure 4A, Supplemental Table 1A).

**Figure 4.**
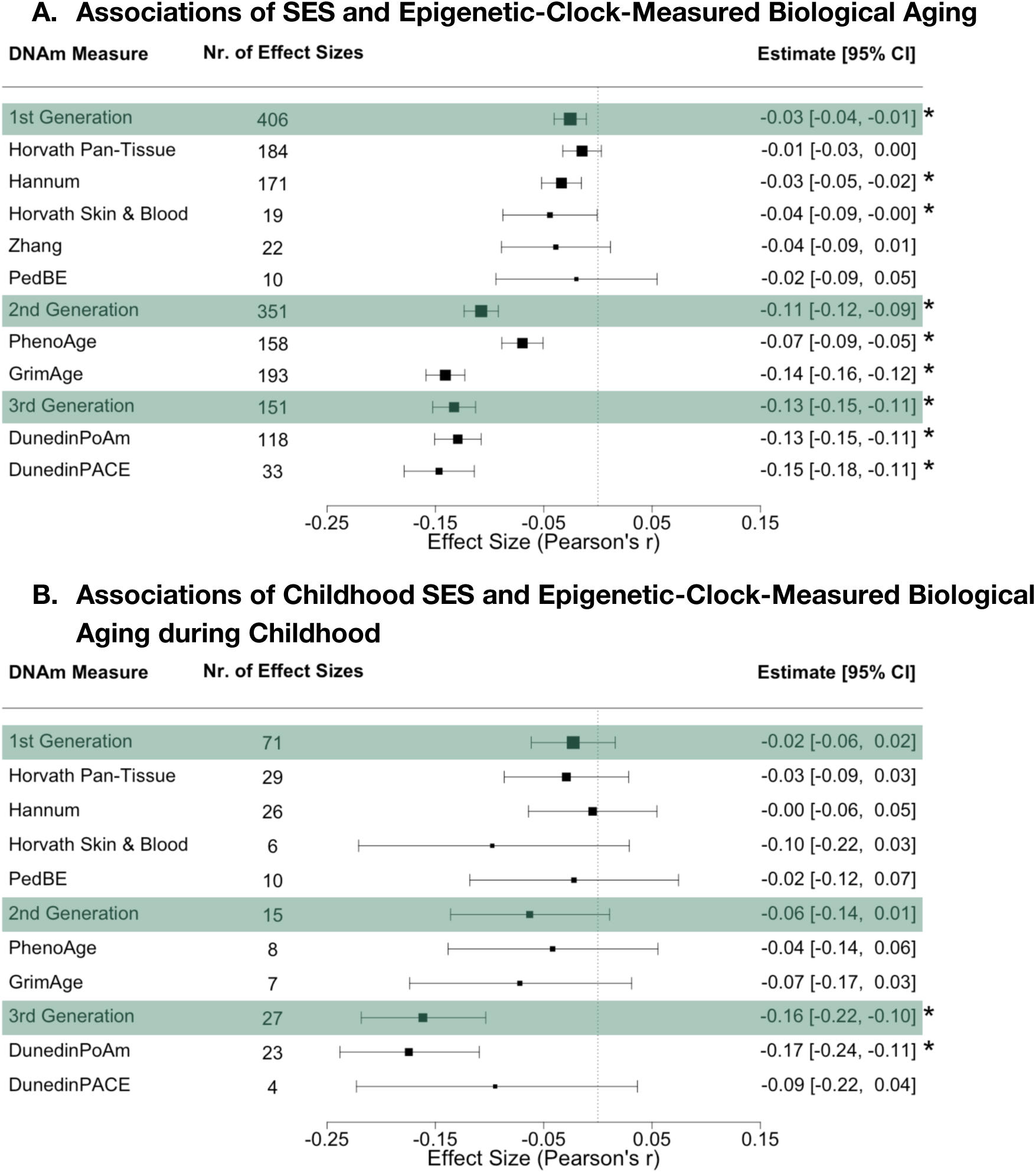
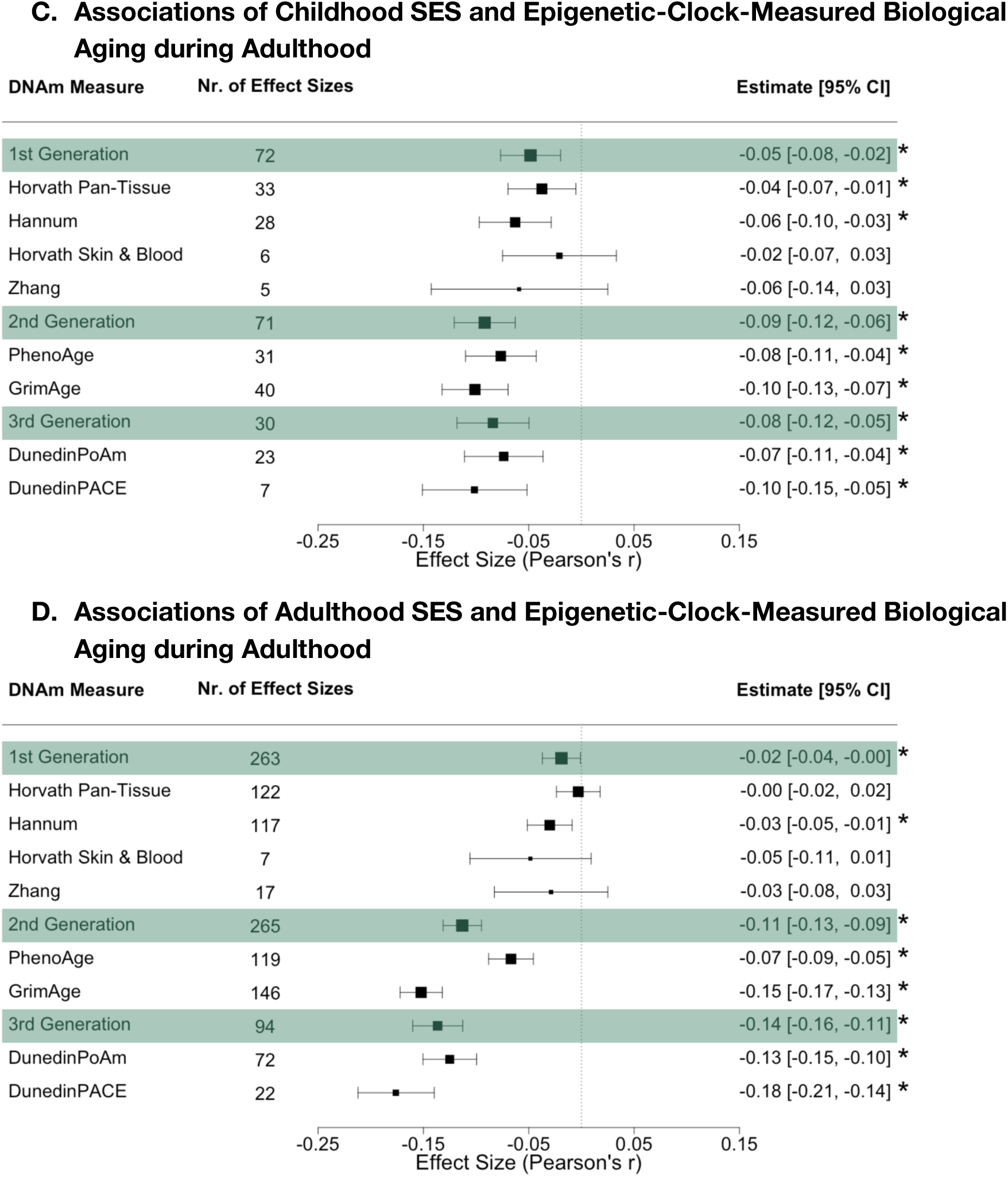
Meta-analytic association between socioeconomic status (SES) and epigenetic-clock-measured biological aging. Forest plots depict meta-analytic associations of **A.** socioeconomic status (SES) and epigenetic-clock-measured biological aging, **B.** childhood SES and epigenetic-clock-measured biological aging measured in childhood. **C.** childhood SES and epigenetic-clock-measured biological aging in adulthood, **D.** adulthood SES and epigenetic-clock-measured biological aging measured in adulthood. Green panels indicate the meta-analyzed effect size estimates for first-, second-, and third-generation clocks, followed by individual epigenetic clocks within each category. Analyses were conducted using Fisher-Z transformation and later transformed to Pearson’s r to ease interpretation. Effect size estimates are bounded by 95% confidence intervals (CIs). * denotes significant difference from zero at p < 0.05. The size of the boxes in the middle panel represents the precision of the effect size estimate (i.e., the smaller the CI, the more precise the estimate).

Among the individual clock measures, the SES-clock associations were most pronounced for GrimAge acceleration, DunedinPoAm, and DunedinPACE compared to all other DNAm clocks (Figure 4A, Supplemental Table 1A). These three measures indicated accelerated and faster aging in lower relative to higher SES individuals (*r* ‘s range from −0.13 to −0.15).

We next conducted non-preregistered sensitivity analyses probing the degree to which SES-clock associations were influenced by findings from the US Health and Retirement Study (HRS), which was the source of the largest number of effect estimates (227 from 1,065 effect sizes). We repeated our analysis excluding HRS effect sizes. Our main results were unchanged: the largest effect sizes were observed for third-generation clocks, then second-generation, and then first, with GrimAge, DunedinPoAm, and DunedinPACE having the largest effect sizes in individual-clock analysis (Supplemental Figure 2, Supplemental Table 1B). We ran further non-preregistered sensitivity analyses to investigate if results differed after removing effect sizes obtained from the three meta-analyses included in our database. Again, the main results were unchanged (Supplemental Figure 3, Supplemental Table 1C).

We then investigated whether publication bias influenced the effect sizes describing the relationship between SES-clock associations. We employed a funnel plot ^29^, followed by Egger’s test for plot asymmetry, which provided information on the possibility of publication bias affecting the results of this meta-analysis ^30^. We found no evidence for publication bias in SES-clock associations (Figure 5, Supplemental Table 1D).

**Figure 5.**
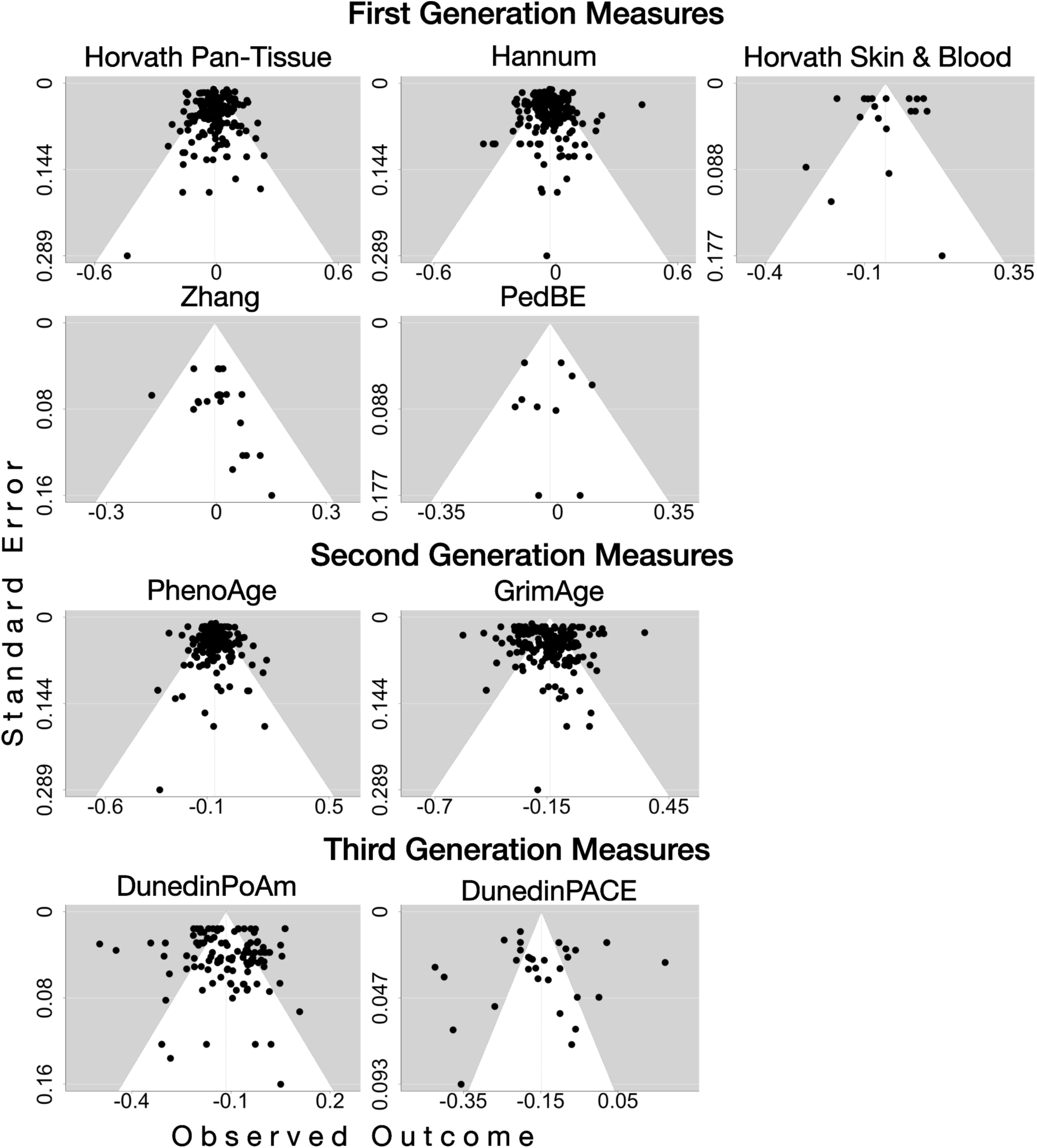
Funnel plots to assess publication bias in the relationship between socioeconomic status and epigenetic-clock-measured biological aging. These plots display individual effect sizes on the x-axis and their standard errors on the y-axis. According to the results of Egger’s regression test, there is no evidence of publication bias for any of the epigenetic-clock-measured biological aging (all ps < 0.05; Supplemental Table 5A).

### (2) Measurement timing effects of SES and epigenetic-clock-measured biological aging

We then probed *when* associations of SES with epigenetic-clock-measured biological aging were apparent. Because of the hypothesized importance of childhood environments on lifelong aging trajectories^31,32^, our main focus was on three combinations of measurement timing: a) children’s DNAm with childhood SES, b) adults’ DNAm with childhood SES, and c) adults’ DNAm with adulthood SES.

We note that DNAm collected in adulthood (83% of effect sizes) is more common than DNAm in childhood (15%). None of the studies had DNAm from the same people collected repeatedly in both childhood and adulthood. Moreover, most cohort studies include either children or adults, but not both. Thus, these analyses can be understood as probing for age and generational cohort differences alongside other study-specific effects (*e.g.,* child studies are substantially more likely to have saliva rather than blood DNAm). First, we split our dataset by DNAm and SES measurement timing to probe measurement timing combinations

### Children’s DNAm with childhood SES

We wanted to know whether children living in families of lower SES compared to higher SES differed in biological aging as measured in child DNAm. Out of the 113 effect sizes, most of the studies came from the USA (60 effect sizes) and UK (34 effect sizes), with a mean age of 10.07 years, ranging from 0-18 years. 37% of the effect sizes were estimated in samples with saliva DNAm and 53% in samples with venous blood.

In an analysis restricted to child DNAm, we found that children living in lower SES families tend to have a faster pace of aging than their more affluent peers. The magnitude of this association between SES and biological aging was moderated by clock generation (*F*(2) = 18.91, *p* < .001) and clock measure (*F*(7) = 22.01, *p* < 0.001, Supplemental Table 2A). We found that only third-generation clocks were significantly associated with SES (Figure 4B; *r* = −0.16 [−0.22, −0.1]; Supplemental Table 2A). Amongst individual clock measures, only DunedinPoAm (*r* = −0.18 [−0.24, −0.11]) was significantly associated with SES. There were not enough effect sizes to compare DunedinPoAm to DunedinPACE.

### Adult’s DNAm with childhood SES

Then we examined whether adults who - as children - grew up in families of lower SES compared to higher SES differed in biological aging as measured in their adult DNAm. Most of the studies had retrospective self-reports of childhood SES. Out of the 173 effect sizes across 6 countries, most came from the USA (145), with a mean age of 56.7 years, ranging from 20-79, and venous blood DNAm was the most common tissue, which was measured in 84% of effect sizes.

In an analysis restricted to childhood SES and adulthood DNAm, we found that adults who grew up in lower SES families tend to have an accelerated biological age and faster pace of aging than their more affluent peers. The magnitude of the association between SES and biological aging was moderated by clock generation (*F*(2) = 14.74, *p* <.001) and clock measure (*F*(7) = 21.71, *p* < .001, Supplemental Table 2B). It was strongest for second- (*r* = −0.09 [−0.12, −0.06]) and third-generation clocks (*r* = −0.08 [−0.12, −0.05), and weakest for first-generation clocks (*r* = −0.05 [−0.08, −0.02]; see Figure 4C; Supplemental Table 2B). Among the individual clock measures, the SES-clock associations were most pronounced for GrimAge acceleration, DunedinPACE, PhenoAge acceleration, and DunedinPoAm (*r’s* range from −0.07 to −0.11, see Figure 4C, Supplemental Table 2B).

### Adult’s DNAm with adulthood SES

Next, we tested if adults currently of lower SES compared to higher SES differed in biological aging as measured in their adult DNAm. Most of the 622 effect sizes came from the USA (427 effect sizes), followed by the UK (42 effect sizes). The mean age was 55.75, ranging from 19.3 to 85.8. The most commonly used tissue was venous blood, contributing 551 effect sizes (89%).

In an analysis restricted to adulthood SES and adulthood DNAm, we found that adults currently of lower SES tend to have an accelerated biological age and faster pace of aging than those of higher SES. The magnitude of the association between SES and biological aging was moderated by clock generation (*F*(2) = 168.98, *p* < .001) and clock measure (*F*(7) = 275.02, *p* < .001, Supplemental Table 2C). It was most pronounced for third-generation clocks (*r* = −0.14 [−0.16, −0.11]), followed by second-generation clocks (*r* = −0.11 [−0.13, −0.09]), and weakest for first-generation measures (*r* = −0.02 [−0.04, −0.00], see Figure 4D, Supplemental Table 2C). Amongst individual clock measures, GrimAge, DunedinPoAm, and DunedinPACE had the largest effect sizes (*r*’s range from −0.13 to −0.18, Figure 4D, Supplemental Table 2C) compared to all other clocks.

### Child and adult DNAm with SES

Then we examined if epigenetic-clock associations with SES significantly differed by measurement timing. We restricted this analysis to GrimAge and DunedinPoAm. For both clocks, the magnitude of the SES association was moderated by measurement timing (DunedinPoAm (*F*(2) = 16,75, *p* <.001; GrimAge (*F*(2) = 14,92, *p* <.001, Supplemental Table 2D). The DunedinPoAm-SES association was significantly weaker when SES was measured in childhood and DNAm in adulthood compared to when DunedinPoAm-SES were both measured in childhood or both measured in adulthood. The DunedinPoAm-SES association was not statistically different when comparing childhood SES with child DNAm to adulthood SES with adult DNAm (Supplemental Table 2D).

The GrimAge-SES association was significantly weaker when SES was measured in childhood and DNAm in adulthood compared to when GrimAge-SES were both measured in adulthood (see Supplemental Table 2D). Although the GrimAge-SES association was not significantly different from zero in child DNAm (*r* = –0.07, [–0.02, 0.18]), the effect size did not significantly differ from the effect size of SES in adulthood with adults DNAm (*r* = –0.15 [–0.17, – 0.13], Supplemental Table 2D). This may be attributed to the wide confidence interval and larger standard error in the child DNAm estimate, which are due to the limited number of effect sizes available for SES–GrimAge associations (see Figure 4B).

As sensitivity analyses, we further delineated whether associations between childhood SES and GrimAge and DunedinPoAm differed when DNAm was measured in “childhood (5 – 10 years)” or “adolescence (10-18 years)”. Moderation analyses showed no significant differences across these age bins (see Supplemental Table 2E). Next, we probed whether the association between adulthood SES and GrimAge and DunedinPoAm differed when DNAm was measured in “young adulthood (ages 18-45)”, “middle adulthood (ages 45-65)”, and “old adulthood (age 65+)”. Moderation analyses showed no significant differences across these age bins (see Supplemental Table 2E).

### Intergenerational SES, intergenerational mobility, and repeated measures of biological aging

Several studies examined clock associations with intergenerational SES, primarily by comparing first- and second-generation clock measures in individuals who had stably low compared to stably high SES across their own and their parents’ generations. We found that individuals whose parents and themselves grew up in low SES had accelerated biological aging compared to individuals whose parents and themselves grew up in high SES (Supplemental Figure 4).

A few studies also investigated intergenerational mobility, primarily by comparing second- and third-generation clock measures in individuals who had higher SES than their parents with individuals who remained in low SES compared to their parents. These studies reported a slower pace of aging in individuals who achieved upward intergenerational mobility compared to those who remained low in SES ^25,33^, but did not yield enough effect sizes to synthesize in a meta-analysis.

Moreover, a few effect sizes explored the association between SES and repeated measures of biological aging within individuals, or between repeated measures of both SES and biological aging within individuals. Meta-analyzing these associations showed no significant link between change in SES and change in biological aging. Considering this was based on few effect sizes, more research is needed to better quantify these associations (see Supplemental Figure 5).

### (3) Differences between race and ethnic groups in biological aging

Next, we examined whether epigenetic-clock-measured biological aging differed between race and ethnic groups, considering the influence of clock generation and measure. All effect sizes included in this analysis come from studies of USA-based cohorts. USA social disparities in health and mortality are most pronounced between Black and White individuals ^34–36^. In our analysis, we found that Black individuals tended to exhibit accelerated and faster biological aging compared to White individuals. The magnitude of this difference between racial groups in epigenetic-clock-measured biological aging was moderated by clock generation (*F*(2) = 88.40, *p* < .001) and measure (*F*(7) = 121.41, *p* < .001, see Supplemental Table 3A). The difference was largest for third-generation measures (*d* = 0.41, 95% CI = [0.28, 0.53]) compared to second-generation measures (*d* = 0.29, CI = [0.18, 0.4]), both of which were larger than first-generation measures (*d* = −0.05, CI = [−0.16, 0.05]) (see Figure 6A; Supplemental Table 3A).

**Figure 6.**
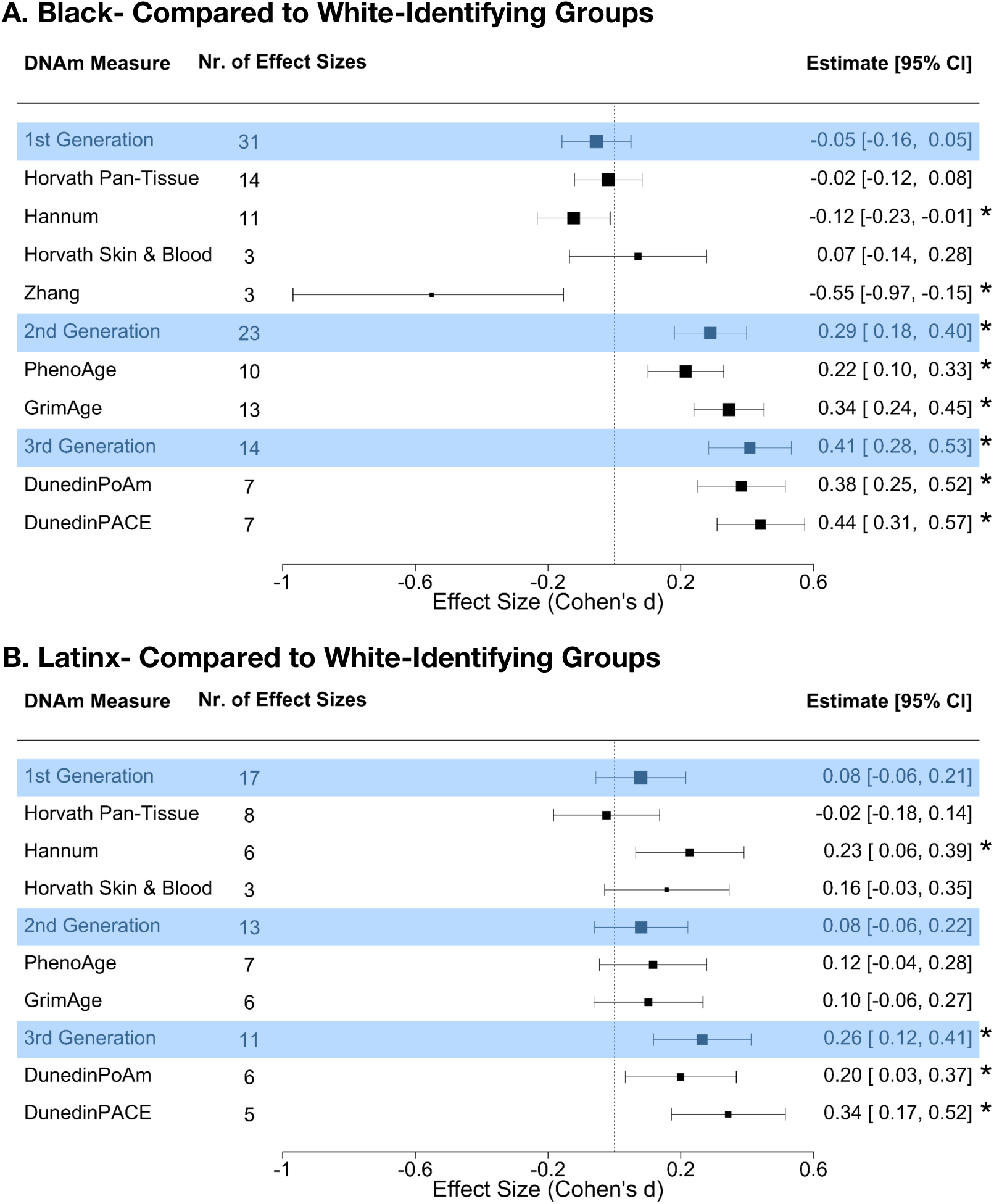
Meta-analytic effect sizes describing the differences between self-Identified race and ethnicity in epigenetic-clock-measured biological aging. Forest plots depict meta-analytic associations of **A.** Black-compared to White-identifying individuals and **B.** Latinx-compared to White-identifying individuals. Green panels indicate the meta-analyzed effect size estimates for first-, second-, and third-generation clocks, followed by individual clock measures within each category. Effect size estimates and confidence intervals (CIs) are reported in Cohen’s *d*. Effect sizes report differences for self-identified racial/ethnic groups in reference to White. **\*** denotes significant difference from 0 at *p <* 0.05. The size of the boxes in the middle panel represents the precision of the effect size estimate (*i.e.,* the smaller the CI, the more precise the estimate).

Among the individual clock measures, the difference between Black- and White individuals was most pronounced in GrimAge acceleration, DunedinPoAm, and DunedinPACE (range of *d’s* 0.34 to 0.44) compared to all other DNAm measures (see Figure 6A; Supplemental Table 3A). These three measures indicated accelerated and faster aging in Black-relative to White individuals. In contrast, the Hannum (*d* = −0.12 [−0.23, −0.01]) and Zhang (*d* = −0.55 [−0.97, −0.15]) clocks indicated biological age *deceleration* in Black-relative to White-individuals.

Second, we examined if epigenetic-clock-measured biological aging differed between Latinx- and White individuals, given social disparities in health between these groups ^37^. We found that Latinx-compared to White individuals tended to exhibit accelerated and faster biological aging. Again, the magnitude of this difference between ethnic groups and DNAm measures was moderated by clock generation (*F*(2) = 14.91, *p* = <.001) and measure (*F*(6) = 71.18, *p* < .001, see Supplemental Table 3B). The difference was largest for third-generation measures (*d* = 0.26 [0.12, 0.41]) compared to second-generation and first-generation measures, both of which were not significantly different from zero (see Figure 6B, Supplemental Table 3B). Among the individual clock measures, the difference between Latinx- and White groups was most pronounced in DunedinPACE (*d* = 0.34 [0.17, 0.52]) compared to all other DNAm measures (see Figure 6B, Supplemental Table 3B, and Supplemental Table 3C for estimates in Pearson’s *r*).

We could not investigate the heterogeneity in race/ethnicity associations with epigenetic clocks by the timing of DNAm measurement, as there were relatively few studies involving racially marginalized children. We did not perform publication bias analyses for race/ethnicity results, because of the limited number of effect sizes and the high level of heterogeneity (at least 80%) in clock differences between groups, which can bias Egger’s tests ^38,39^.

### (4) Sex

We explored whether the strength of SES-clock associations varied depending on sex. Few studies included SES-clock associations for females and males separately. In line with earlier meta-analyses ^40,41^, we did this by comparing associations of sex ratio balanced and sex ratio imbalanced (>65%females or males). We found no moderating effects of sex for SES-GrimAge acceleration and SES-DunedinPoAm associations (see Supplemental Table 4A). There were not enough effect sizes to probe the sex moderation of SES-DunedinPACE associations. There were no moderating effects of sex for other epigenetic clocks, except for Hannum and PhenoAge, where associations were more pronounced when derived from predominantly female compared to sex ratio balanced samples (see Supplemental Table 4B).

We did not probe whether race/ethnicity differences with epigenetic clocks differed by sex composition of the sample, because there were too few studies with sex-imbalanced cohorts.

### (5) Technical factors

We examined whether the strength of SES-clock associations varied depending on technical factors (e.g., tissue type, array, cell correction). However, our analysis was substantially hindered by inconsistent or missing reporting of which technical factors were included as covariates. Thus, our findings should be considered preliminary and require more examination in future studies.

First, we examined if SES-clock associations were moderated by tissue type (e.g., blood, saliva, buccal DNAm). We found no moderating effect of tissue type (see Supplemental Table 4A). We note that there were not enough effect sizes to probe tissue type moderation of SES-DunedinPACE associations. Next, we explored whether associations varied depending on the array type (EPIC, 450k). We found no differences in SES-clock associations between arrays for clock generation or individual clocks (Supplemental Figure 6; Supplemental Table 5A-B).

Then we probed if the strength of SES-clock associations varied depending on whether studies applied statistical corrections for cell composition. Our results indicated that in venous blood samples, cell composition correction moderated the SES-Hannum acceleration association, such that the association was stronger when effects were not corrected for cell-type variation (Supplemental Table 5C). In saliva samples, cell composition correction moderated the SES-Hannum and SES-Horvath Pan-Tissue acceleration association, such that the association was stronger when effects were not corrected for cell-type variation (Supplemental Table 5D). There were not enough effect sizes to run this analysis for other saliva DNAm measures.

Only analysis of array and cell correction had enough effect sizes to examine the moderation of race/ethnicity differences with epigenetic clocks. We found no moderation effects (see Supplemental Table 5E).

## Discussion

We conducted a pre-registered, multi-level meta-analysis synthesizing 1,065 effect sizes across 140 studies on the associations of SES or race/ethnicity with three generations of epigenetic-clock measures of biological aging. The included studies comprised N = 65,919 people from 23 countries aged 0 to 86 years old. There were four key findings.

*First*, individuals of lower SES tended to appear biologically older and to be aging faster compared to individuals of higher SES. When considered by generation, effect sizes were most consistent for the third generation of epigenetic clocks, known as epigenetic speedometers, which were developed by modeling Pace of Aging, a composite phenotype of summarizing rates of change across multiple organ systems. These were followed by the second generation of epigenetic clocks, which were developed by modeling mortality risk. In contrast, the first generation of epigenetic clocks, which were developed by modeling chronological age, demonstrated small or non-significant associations with SES. When considered at the level of individual epigenetic clocks, third-generation DunedinPoAm and DunedinPACE and second-generation GrimAge exhibited the strongest associations with SES. Strikingly, we did not find evidence of publication bias in SES-clock associations. This builds confidence in the meta-analytic effect-size estimates we report ^42^.

*Second*, the strength of SES associations with third-generation clocks was similar in magnitude when DNAm is measured in childhood (<18 years) compared to adulthood, whereas SES associations with second-generation clocks were weaker in studies of children as compared with studies of adults. Second-generation clocks were included in fewer studies of children as compared with third-generation clocks. As a result, although the effect sizes differed between age groups by a factor of two, the difference did not reach statistical significance. Interestingly, childhood SES showed similar-magnitude associations with second- and third-generation clocks measured from adult blood samples. One interpretation of these data is that acceleration to aging processes caused by early adversity is immediately apparent in “speedometer” type third-generation clocks, but does not become apparent until later in life in second-generation clocks^32,43^. Collectively, findings suggest that third-generation clocks, such as DunedinPACE, are the most promising tools for tracing environmental impacts on aging and health early in life.

*Third*, individuals of socially marginalized racial and ethnic groups show signs of accelerated biological aging relative to White-identifying individuals. The fact that all effect sizes available for this analysis came from US-based studies highlights the need for epidemiological research on this topic in other countries. In line with observations about disparities in social conditions and in health and mortality^34–37^, group differences were larger for Black-White comparisons than for Latinx-White comparisons. Similar to our findings for SES, those racial and ethnic disparities were most pronounced for third-generation epigenetic clocks, followed by second-generation clocks. Again, DunedinPoAm, DunedinPACE, and GrimAge showed the starkest social differences in epigenetic-clock-measured aging. We did not perform publication bias analyses for race/ethnicity results, because of the limited number of effect sizes and the high level of heterogeneity.

Racism intersects with socioeconomic disadvantage in addition to other health-damaging factors, creating a complex web of challenges ^44–46^. Consistent with this interpretation, effect sizes were somewhat larger for race/ethnic disparities as compared with SES disparities (*e.g.,* DunedinPACE *r* with SES = −0.15 vs Black-White *r* = 0.22). However, because current studies have relied on self-reported race and ethnicity, they lack the nuances of probing structural (*e.g.,* neighborhood segregation) and individual-level (*e.g.,* personal experiences of discrimination) processes of how racism may affect biological aging ^23^. Moreover, we could not investigate the heterogeneity in racial and ethnic differences in epigenetic clocks by the timing of DNAm measurement, as there were relatively few studies involving racially marginalized children. Recent research in over n=2000 individuals from the Future Families and Child Well-Being cohort find that these racial and ethnic differences are visible in 9-year-old’s saliva GrimAge, PhenoAge, and DunedinPACE and become larger in repeated measures of DNAm from age 9 to 15 years in all three metrics ^23,47^.

Fourth, we found little evidence of moderating effects of sex, array, tissue type and cell-count correction on SES associations or racialized differences with epigenetic-clock-measured biological aging. However, we believe the question of whether these factors may influence SES-clock associations requires further investigation. For example, future studies should directly contrast SES-clock associations for females and males to test whether associations are similar for men and women. In this meta-analysis, we were limited to comparing effect sizes across studies that included mostly women, mostly men, or sex-balanced samples. Among adults, men tend to exhibit older biological age and faster pace of aging as compared to women of the same chronological age ^18,24^. In contrast, among adolescents, this pattern is reversed, with girls exhibiting older biological age and faster pace of aging as compared to same-aged boys ^23,48,49^, possibly aligning with earlier pubertal maturation seen in girls relative to boys ^50,51^. Sex-specific racial and ethnic disparities in biological aging are less examined. Our results indicate that despite sex differences in biological aging and health, social disparities in GrimAge and DunedinPACE are comparable across sexes.

Additionally, Future research should prioritize clear reporting regarding the selection of covariates and consider providing effect sizes with and without various covariate adjustments, because our analysis was limited by a lack of information in many studies. Future research should prioritize clear reporting regarding the selection of covariates and consider providing effect sizes with and without various covariate adjustments. For instance, recent studies comparing the similarity of epigenetic clocks across blood, saliva, and buccal DNAm collected from the same people highlight that epigenetic clocks do differ between cell types, especially in terms of absolute values compared to between-person differences, and the degree of this cross-tissue correspondence differs by clock measure ^52,53^. Hence, our findings indicating that SES associations with GrimAge acceleration, DunedinPoAm, and DunedinPACE are robust to variation in technical factors should be treated with caution.

These findings should be interpreted within the framework of several limitations. Our meta-analysis was restricted to studies preprinted or published before February 2024. Since then, several other studies have examined associations of SES or racial and ethnic disparities in epigenetic clocks mirroring our results ^54,55^. In addition, there are epigenetic clocks not included in our analysis, including new clocks for which studies of SES have not yet been conducted^56,57^. Our analyses of sex differences and technical modifiers affecting SES-clock associations lacked complete information. Few studies reported formal tests of sex differences. Some studies failed to report complete information on technical details. And the number of studies conducted in non-blood tissues remains small. These questions should be revisited as more published data are available. Finally, analysis of race/ethnic variation in epigenetic clocks may be confounded by ancestry-linked genetic artifacts. DNA-based research overrepresents European ancestry populations^58,59^, which could create artifacts in DNAm datasets that include ancestrally more distal groups^60^. However, there is little evidence to date to support this type of bias in the case of the second- and third-generation clocks; studies find consistent patterns of clock associations with healthspan and lifespan within different race/ethnic groups and have failed to detect evidence of ancestry-linked genetic variation in clock values ^25,61–65^. Regardless, there remains an urgent need to develop biological aging algorithms that are inclusive and reflective of aging processes in diverse populations.

In conclusion, our meta-analysis found evidence that low SES and marginalized race/ethnicity are associated with accelerated biological aging as measured by epigenetic clocks. Third-generation “epigenetic speedometers” and the second-generation GrimAge clock showed the strongest and most consistent associations in adults. In children, only the third-generation speedometers showed consistent associations. Our findings provide guidance for the selection of epigenetic clocks for studies of social determinants of health.

There is now abundant correlational evidence linking a range of socially stratified exposures with epigenetic clock measures of biological aging, including environmental toxicants, access to healthcare, stress, nutritious food, opportunities for exercise and quality sleep ^3,7,66–69^. The next phase of research must advance knowledge from simple correlations toward the identification of causal relationships linking social disparities to biological aging. One path forward is through analysis of samples collected in intervention trials. For example, epigenetic clock measures of aging were modified by changes in behavior and nutrition ^70,71^. The same approach can be applied to policy trials or other interventions Our results can inform these studies in their selection of outcome measures.

## Methods

The Preferred Reporting Items for Systematic Reviews and Meta-Analyses (PRISMA) Checklist^73^ was used to structure, conduct and report this meta-analysis.

### Search Terms

Table 1 presents an overview of the variables of interest for Epigenetic-clock-measured biological aging and social determinants of health, which we restrict to measures of socioeconomic status (SES) and racialization. These variables were used to produce a search string for a systematic literature search (see *Search Strategy and Information Sources*).

### Search Strategy and Information Sources

For the literature search, the following databases were used: PubMed, PsycINFO, Web of Science, medRxiv, and bioRxiv. Search for medRxiv and bioRxiv was performed using R’s *medrxivr* package ^78^. Literature search was completed at one time point, and not repeated thereafter. The search string was used to scan titles and abstracts, with specific search terms listed in Supplemental Table 6. We included studies published or pre-printed between 2013 (when the first epigenetic clock was published, the Horvath pan-tissue clock) and February 2024 (when the meta-analytic search was conducted).

### Eligibility Criteria

Study inclusion and exclusion criteria are presented in Table 2.

**Table 2.** Inclusion and exclusion criteria for this meta-analysis.

| Inclusion Criteria | Exclusion Criteria |
| --- | --- |
| - Written in English<br>- Publishing date after 2013 (first publication of Epigenetic-clock-measured biological aging)<br>- Reporting effect size(s) including the association between at least one social determinant of health and one DNAm measure of biological aging | - Reviews, dissertations, book chapters, conference abstracts<br>- Studies on clinical populations<br>- Age acceleration not reported for first- and second- generation measures<br>- Effect sizes not reported and not provided by authors upon request |

### Selection Procedure

Zotero was used to manage and organize papers. Authors ADR and AB independently screened the obtained articles based on their titles and abstracts using ASReview^78^, an open-source AI-assisted program for systematic reviews. Then, selected and rejected articles were compared and where differences arose, they were resolved through discussion. This step led to the exclusion of 420 articles. The two authors then independently performed full-text screening of the remaining 438 articles. After discussion over disagreements, a further 326 articles were excluded (Figure 2).

During the full-text screening process, 114 articles that included relevant SES/race/ethnicity and biological aging measures but did not report the effect sizes of their associations were identified. Emails were sent to the corresponding authors to request the effect sizes. The authors were given three weeks to provide the data. A reminder email was sent two weeks after the initial email as a follow-up. Out of these 114 articles, we did not receive a response for 71 (62.3%) articles. Out of the authors that responded, data were received for 19 (16.7%) articles. 10 articles we did not receive data for still included relevant results, and were therefore included in this meta-analysis.

The references of the articles selected for inclusion were screened by ADR to find relevant articles that were not identified by the initial literature search. Among the 41 articles identified in this step, 19 included relevant SES and biological aging measures but did not report effect sizes. Requests for data were sent via email as previously described. We did not receive any response for 9 (56.3%) articles and data were received for 6 (31.6%) articles. This step led to the inclusion of 28 articles, resulting in a final meta-analytic sample of 140 articles.

### Coding of Studies

A coding scheme was developed based on meta-analysis guidelines ^79^, including study descriptives (*e.g.,* authors, title, year of publication, etc.) and study characteristics (*e.g.,* sample size, country, participants, DNAm measure(s) of biological aging, SES measure, race, effect sizes, etc.). Potential moderator variables of interest (see “Moderator Analyses” section) were listed in the coding scheme. While Epigenetic-clock measures of biological aging were consistently reported as continuous variables, this was not always the case for SES measures. Some studies reported them as continuous variables (e.g., years of education), while others reported them as categorical variables (e.g., highest educational attainment). Effect sizes for associations between two continuous variables were taken as reported. For studies reporting the association between biological aging and categorical variables, only the effect size denoting the difference between the lowest and highest categories was extracted. To ensure consistency, SES variables were adjusted when necessary, so that higher values indicated higher SES.

20% of the articles were randomly selected to be coded by three authors, ADR, MA, and AB. Two methods were used to evaluate inter-rater reliability. For continuous variables, intraclass correlation was calculated and it ranged from 0.85 to 1. For categorical variables, Cohen’s κ was measured, with values between 0.57 and 1. Where disagreements arose, they were resolved by in-depth reading and discussion with author YW. After this interrater step, the remaining studies were divided and coded separately by the raters.

The preferred effect size metric was Pearson’s correlation coefficient *r*. However, if Pearson’s *r* was not reported in a given study, the following effect size metrics were extracted: Cohen’s *d*, standardized regression coefficient *β*, unstandardized regression coefficient *b*, Spearman’s rho, or the *t*-statistic from a regression or *t*-test. These metrics were then converted into Pearson’s *r* using R.

Effect sizes reported as Cohen’s *d* were converted into Pearson’s *r* using the function *d*_*to*_*r*() in the “effectsize” package ^80^. Spearman’s rho was transformed into Pearson’s *r using* the conversion formula ^81^. The *t*-statistic from regression was converted using the *t*_*to*_*r*() function from the “effectsize” package, and the *t*-statistic from a *t*-test was converted using *esc*_*t*() from the “esc” package. The standardized regression coefficient *β* was transformed based on a widely used formula for effect sizes whose absolute value did not exceed 0.5 ^82^. For *β* values above this threshold, the function *esc*_*beta*() from the “esc” package ^83^ was used. To convert the unstandardized regression coefficient *b*, two approaches were employed. Effect sizes from categorical SES variables were converted to Pearson’s *r* using the *esc*_*B*() function from the “esc” package. To convert effect sizes from continuous variables, the unstandardized *b*was standardized by multiplying it by the standard deviation (SD) of the predictor variable (i.e., SES measure) and dividing it by the SD of the outcome variable (i.e., biological age acceleration). This standardized regression coefficient was then converted into Pearson’s *r* as described above.

If SDs of SES or age acceleration measures were not reported in a study, various methods were used to impute the SDs. For studies reporting age acceleration and/or SES measure’s SDs for different groups (e.g., men and women, White- and Black-identifying), the pooled SDs for the whole sample were calculated, assuming equal variances of subgroups. For studies reporting interquartile ranges (IQR), the IQR was transformed into SD by dividing the IQR by 1.35 ^84^, assuming the outcome SD is similar to the normal distribution. For studies reporting means and SDs of income/wealth variables in descriptives, but log-transforming them for the analysis, the SD of the log-transformed data was calculated using the formula reported in ^85^.

If SDs of age acceleration measures were not reported, they were taken from another study using the same cohort. The studies with the highest sample size, and the ones closest in age to the target sample size were prioritized. If this was not available, SDs for adulthood and childhood age acceleration measures were taken from other studies that matched in tissue. For effect sizes reporting on age acceleration measured in adults from blood samples, SDs were taken from Health and Retirement Study. SDs for Horvath Pan-Tissue, Horvath Skin and Blood, Hannum, PhenoAge, GrimAge and DunedinPoAm were taken from ^24^. DunedinPACE was taken from ^18^. SD for Zhang clock was taken from the Multi-Ethnic Study on Atherosclerosis ^86^.

SDs for children’s saliva were imputed using data from the Future of Families and Child Wellbeing Study (previously called Fragile Families and Child Wellbeing Study, FFCW). Methylation data processing is described in the “biomarker Data” file on the FFCW documentation website. SDs were calculated by author YW based on data access ^23^. For the remaining childhood samples based on venous blood, cord blood, or buccal tissue, SDs were taken from other studies matching in tissue. If this was not possible, authors were emailed. Finally, if SDs were not reported and could not be taken from another study, authors were emailed. 6 authors provided relevant information.

Once the Pearson’s *r* values were calculated for each effect size, they were converted into Fisher’s *z* for analyses using the *convert*_*r*2*z*() function from the “esc” package. This was done to eliminate bias due to sampling distributions, as the distribution of Fisher’s *z* approximates a normal distribution ^79^. After the analyses, the *z*-scores were converted back to Pearson’s *r* using the *convert*_*z*2*r*() function from the “esc” package for interpretation.

14 effect sizes as biweight midcorrelation (bicor) estimates. As there is not a direct transformation from bicor to Pearson’s *r*, effect sizes reporting bicor values were meta-analyzed separately and reported in Supplemental Figure 7.

### Statistical Analyses

The analyses were conducted using R’s *metafor* package ^87^. A three-level mixed-effects model was employed to accommodate the dependency of effect sizes within and across studies ^88^, as some studies use the same sample and/or report multiple effect sizes. The study is nested within-sample in order to deal with potentially non-independent effect sizes coming from the same article or the same sample of participants. This allows us to include all effect sizes of included studies, while taking the dependency of studies using data of the same cohort into account. In the three-level meta-analytic approach, level 1 accounts for the sampling variance of effect sizes, level 2 considers the variance among effect sizes within the same sample, and level 3 addresses the variance between studies ^89^. This allows the inclusion of all studies while ensuring that studies reporting multiple effect sizes will not have a greater contribution to the mean effect size than those reporting only one effect size ^9^. These analysis steps have been used in previous meta-analyses including DNAm measures ^90^. This analysis was carried out based on the guidelines by Assink and Wibbelink ^91^.

### Main Analyses

Social determinants of health measures were separated into SES and race/ethnicity. The following steps were performed for SES and each race/ethnicity category. First, we assessed whether the association between SES/race/ethnicity measure and epigenetic-clock-measured biological aging differs for first-, second-, and third-generation clock measures (see Table 2). We then investigated whether the association between SES/race/ethnicity measures and clock measures differs for specific clocks, if there were a minimum of 5 effect sizes for a given clock (as with a small number of studies, statistical power is low ^92^). Then, likelihood-ratio tests were conducted to establish the within- and between-cohort heterogeneity separately for the associations of first-, second-, and third-generation clocks and SES. Significant heterogeneity at these two levels would suggest evidence for variance that cannot be solely attributed to sampling variance ^91^.

### Moderator Analyses

If heterogeneity was observed in effect sizes and if there were a minimum of 5 effect sizes per category, moderator analyses were conducted (we deviated from our preregistration that detailed 5 *studies*, to be more inclusive). The moderator analyses were carried out as follows: If there were enough effect sizes for each individual DNAm measure, we assessed whether the moderators (see Table 3) influenced the association between SES/race/ethnicity and DNAm measures, if there were at least 5 effect sizes reported for a given category of the moderator.

**Table 3.**
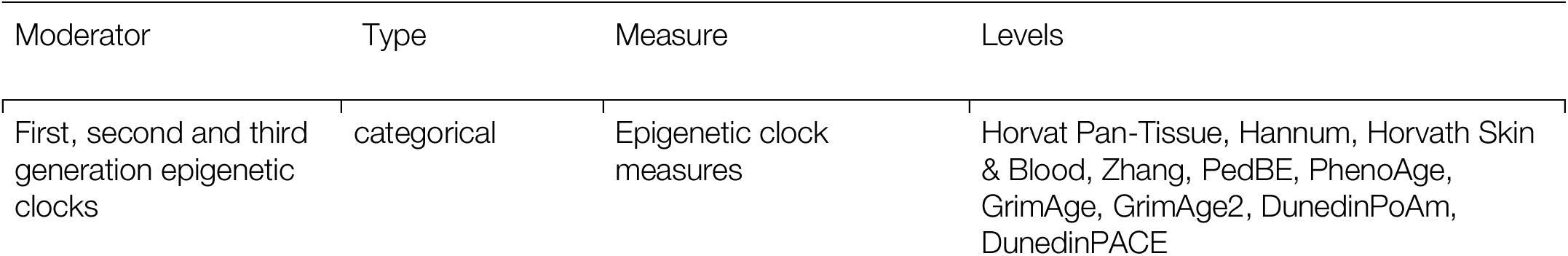
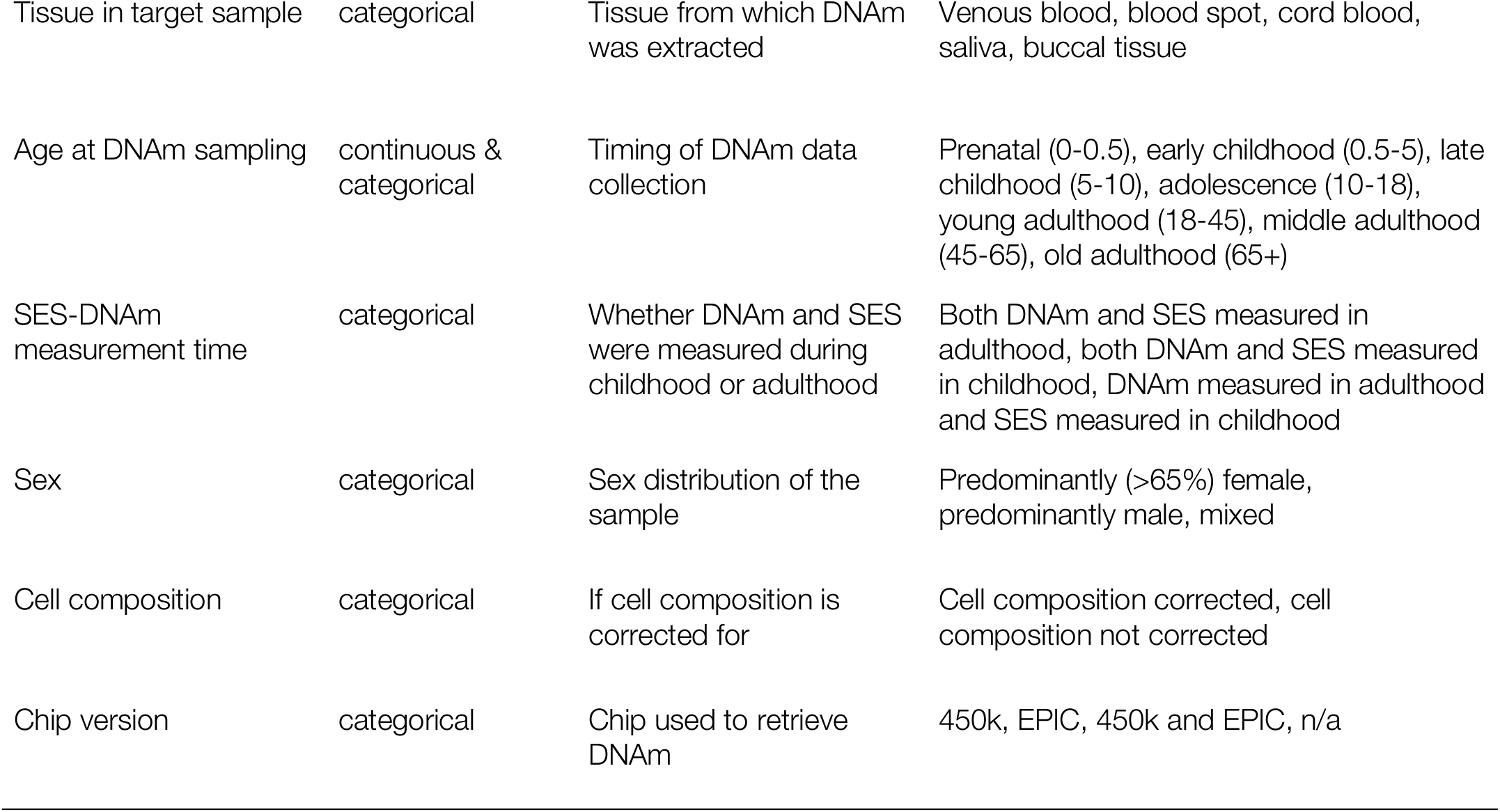
Overview of moderator variables that were explored in this meta-analysis.

## Data Availability

All data produced in the present work are contained in the manuscript

## Acknowledgements

We would like to thank Dr. Wei Perng, Dr. Anna Kankaanpää, Dr. Joanne Ryan, Dr. Zachary Harvanek, Dr. Rajita Sinha, Dr. Maja Popovic, Dr. Thais Lopes de Oliveira, Dr. Sara Hägg, Dr. Joseph Allen, Dr. Daniel McCartney, Dr. Riccardo Marioni, Dr. Tiina Föhr, Dr. Tiffany Powell-Wiley, Julia Nakamura, Dr. Juulia Jylhävä, Abby Hillmann, Dr. Steven Beach, Dr. Chris Verschoor, Dr. May Ahmad Baydoun, Dr. Brooke Gavin McKenna, Dr. Erin Dunn, Dr. Paige Hulls, Dr. Rebecca Richmond, Dr. Jane Maddock, Dr. Erika Wolf, Dr. Natan Yusupov, Dr. Idan Shalev, Dr. Giovanni Fiorito, Dr. Brian Joyce, Dr. Divya Joshi, Dr. Parminder Raina, Dr. Cathal McCrory, Dr. Mariona Bustamante, Dr. Dan Eisenberg, Dr. Larissa Strath, Dr. Yenisel Cruz-Almeida, Dr. Nancy Krieger and Nykesha Johnson for sharing their results with us.

## Supplemental Figures

**Supplemental Figure 1.**
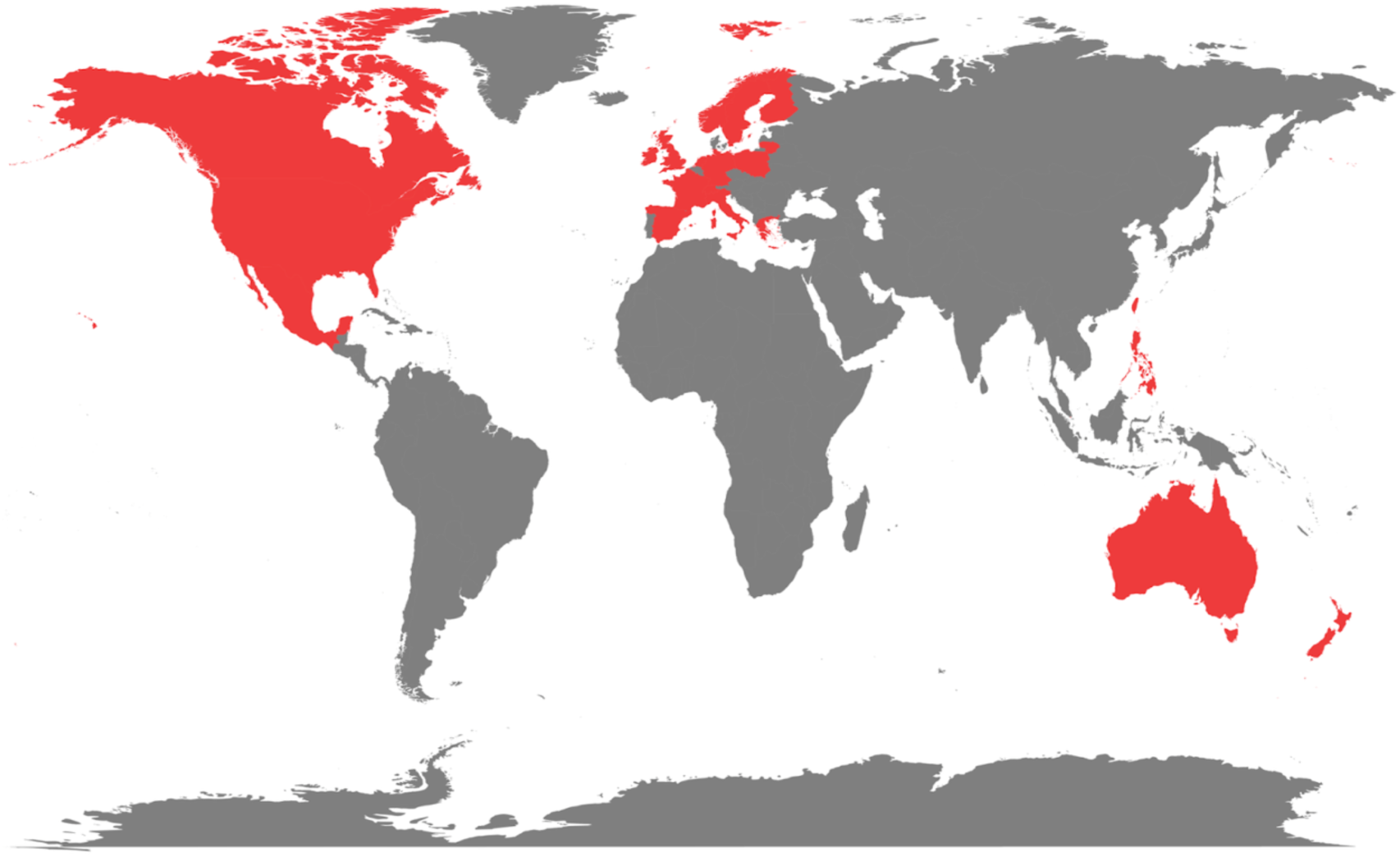
World map showing the countries included in this meta-analysis. Red color indicates that there is at least one effect size from that country (including contributions to meta-analyses). Countries colored in gray does not contribute any effect sizes.

**Supplemental Figure 2.**
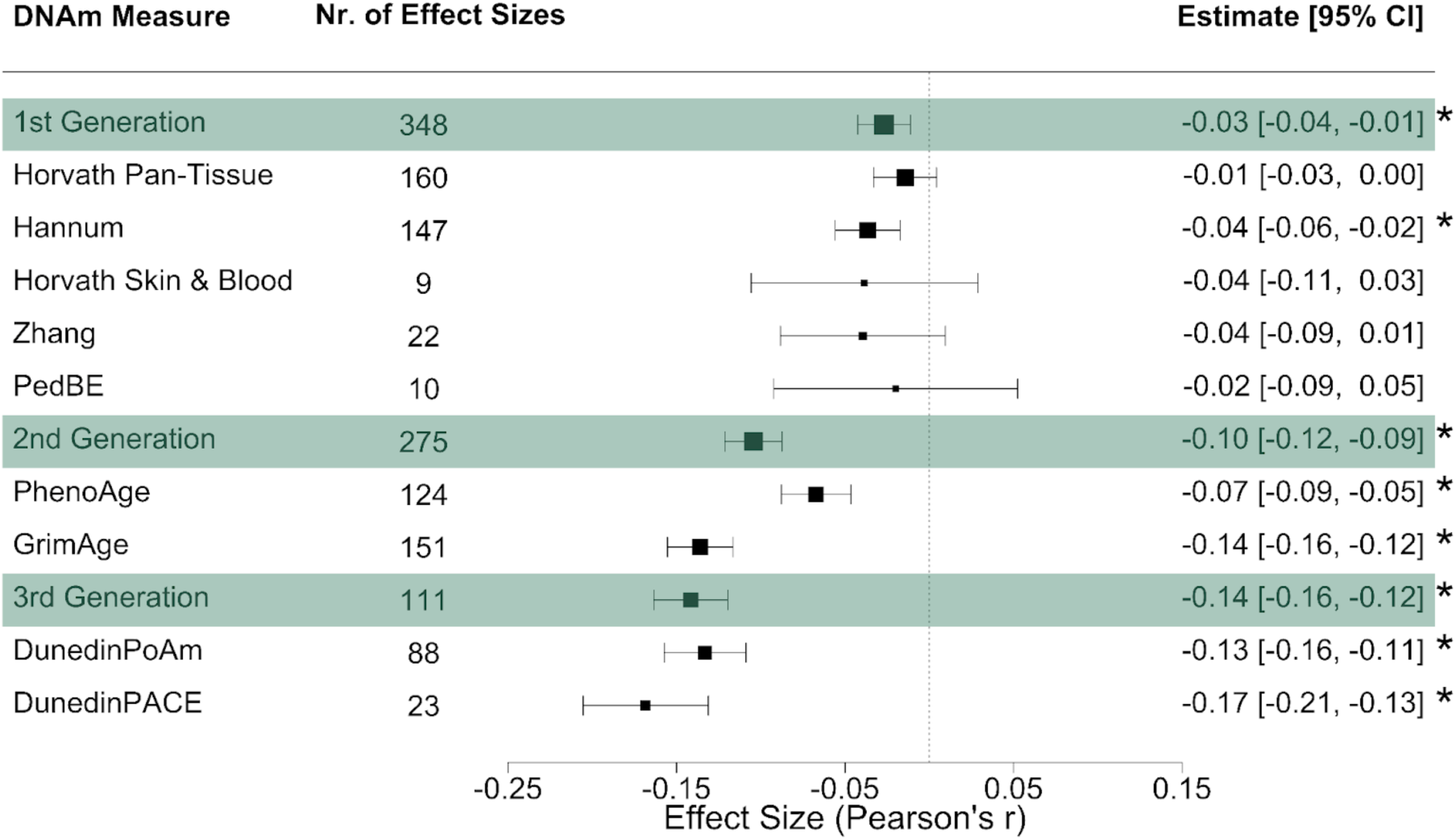
Meta-analytic associations between socioeconomic status (SES) and epigenetic clocks, excluding effect sizes from the Health and Retirement Study (HRS). Forest plot depicts the meta-analytic associations between SES and generations of epigenetic clocks, after 174 effect sizes contributed by HRS are removed. Green panels indicate the meta-analyzed effect size estimates for first-, second-, and third-generation clocks, followed by individual clocks within each category. Analyses are conducted using Fisher-Z transformation and later transformed to Pearson’s r to ease interpretation. Effect size estimates are bounded by 95% confidence intervals (CIs). * denotes significant difference from 0 at p < 0.05. The size of the boxes in the middle panel represents the precision of the effect size estimate (i.e., the smaller the CI, the more precise the estimate).

**Supplemental Figure 3.**
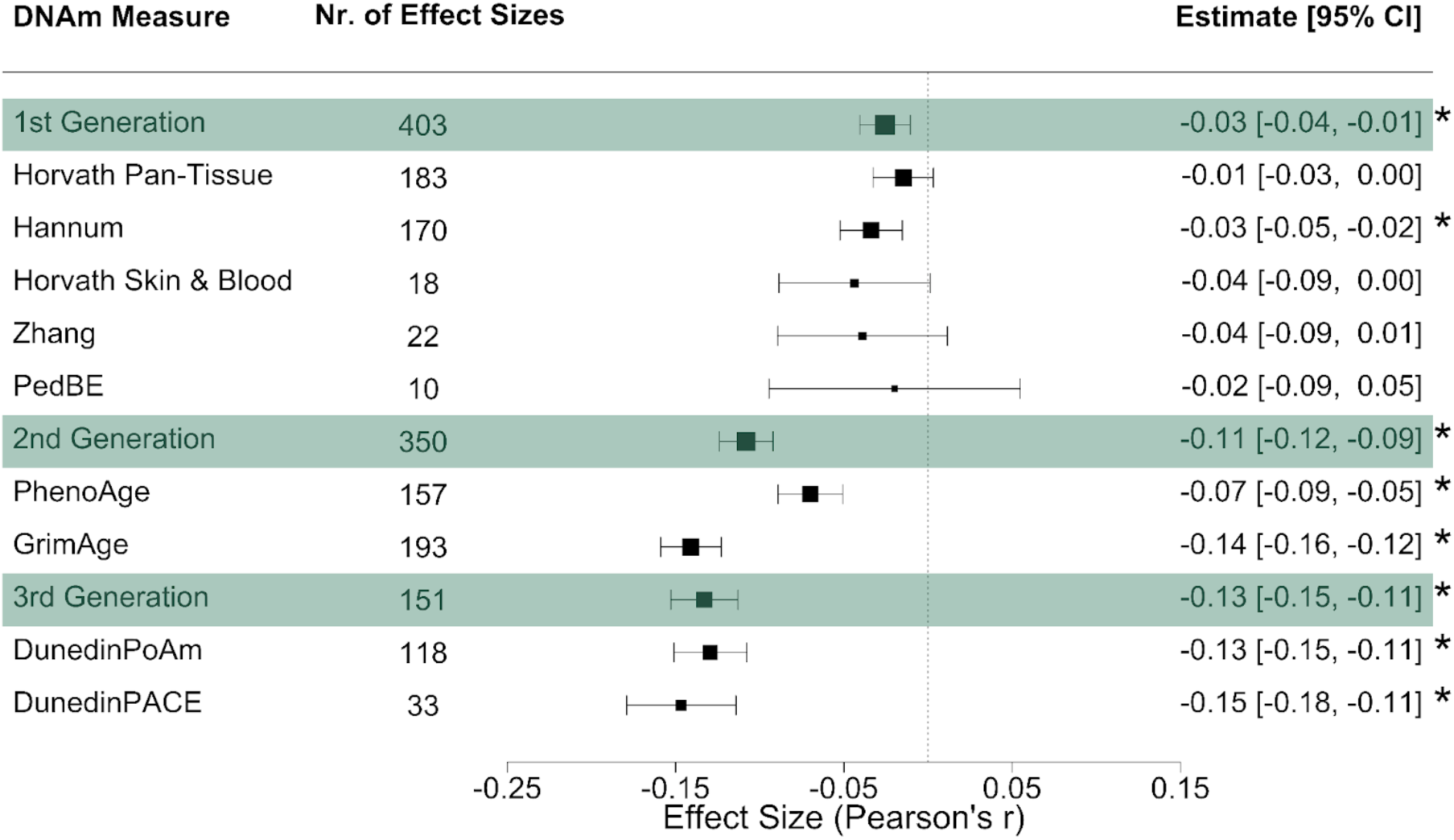
Meta-analytic associations between socioeconomic status (SES) and epigenetic clocks, excluding effect sizes from meta-analyses. Forest plot depicts the meta-analytic associations between SES and generations of epigenetic clocks, after 4 effect sizes contributed by meta-analyses are removed. Green panels indicate the meta-analyzed effect size estimates for first-, second-, and third-generation clocks, followed by individual clocks within each category. Analyses are conducted using Fisher-Z transformation and later transformed to Pearson’s r to ease interpretation. Effect size estimates are bounded by 95% confidence intervals (CIs). * denotes significant difference from 0 at p < 0.05. The size of the boxes in the middle panel represents the precision of the effect size estimate (i.e., the smaller the CI, the more precise the estimate).

**Supplemental Figure 4.**
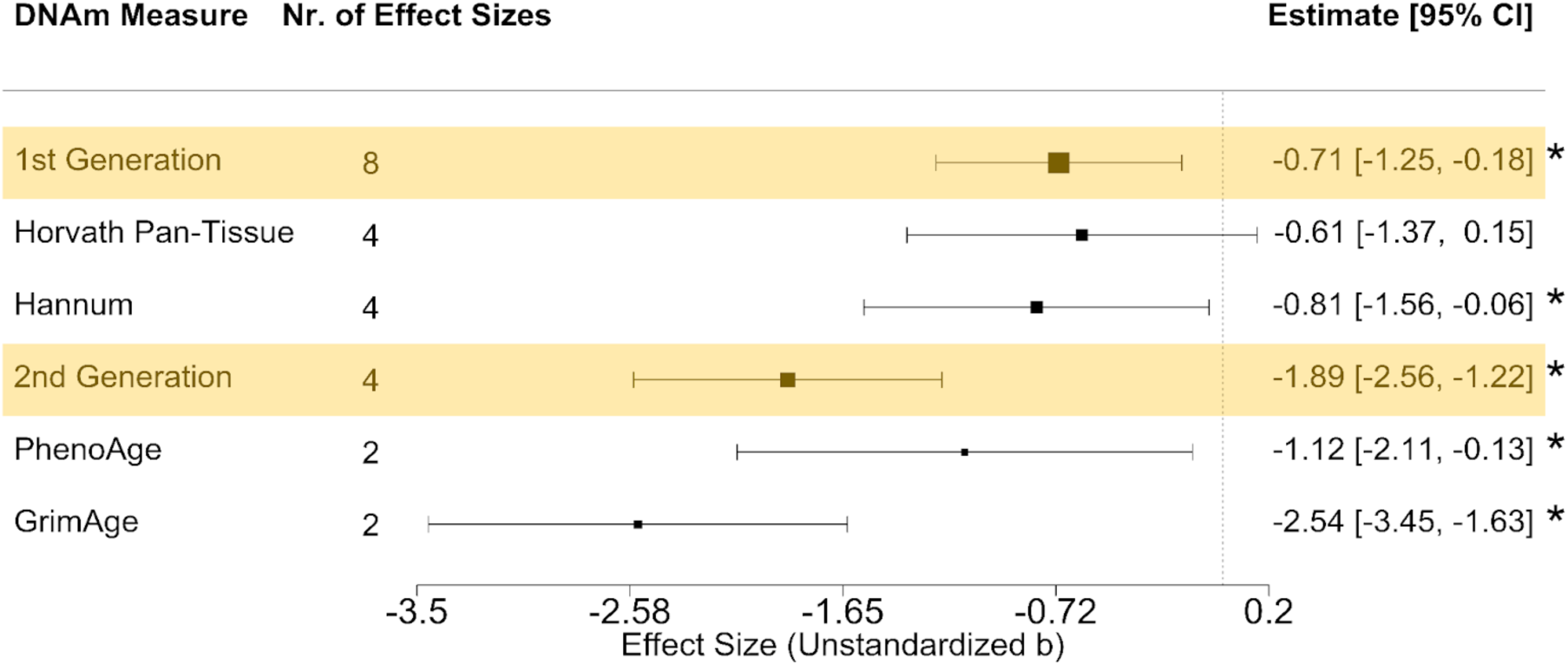
Meta-analytic associations between intergenerational socioeconomic status (SES) and epigenetic clocks. Forest plot depicts the meta-analytic associations between intergenerational SES and epigenetic clocks. DNAm was measured at one time point. Intergenerational SES is measured by the intergenerational stability in SES, comparing parental occupation with current occupation, where the reference group is stably low SES and the target group is stably high SES. Yellow panels indicate the meta-analyzed effect size estimates for first- and second-generation clocks, followed by individual clocks within each category. Results are meta-analyzed as unstandardized regression coefficients. Effect size estimates are bounded by 95% confidence intervals (CIs). * denotes significant difference from 0 at p < 0.05. The size of the boxes in the middle panel represents the precision of the effect size estimate (i.e., the smaller the CI, the more precise the estimate).

**Supplemental Figure 5.**
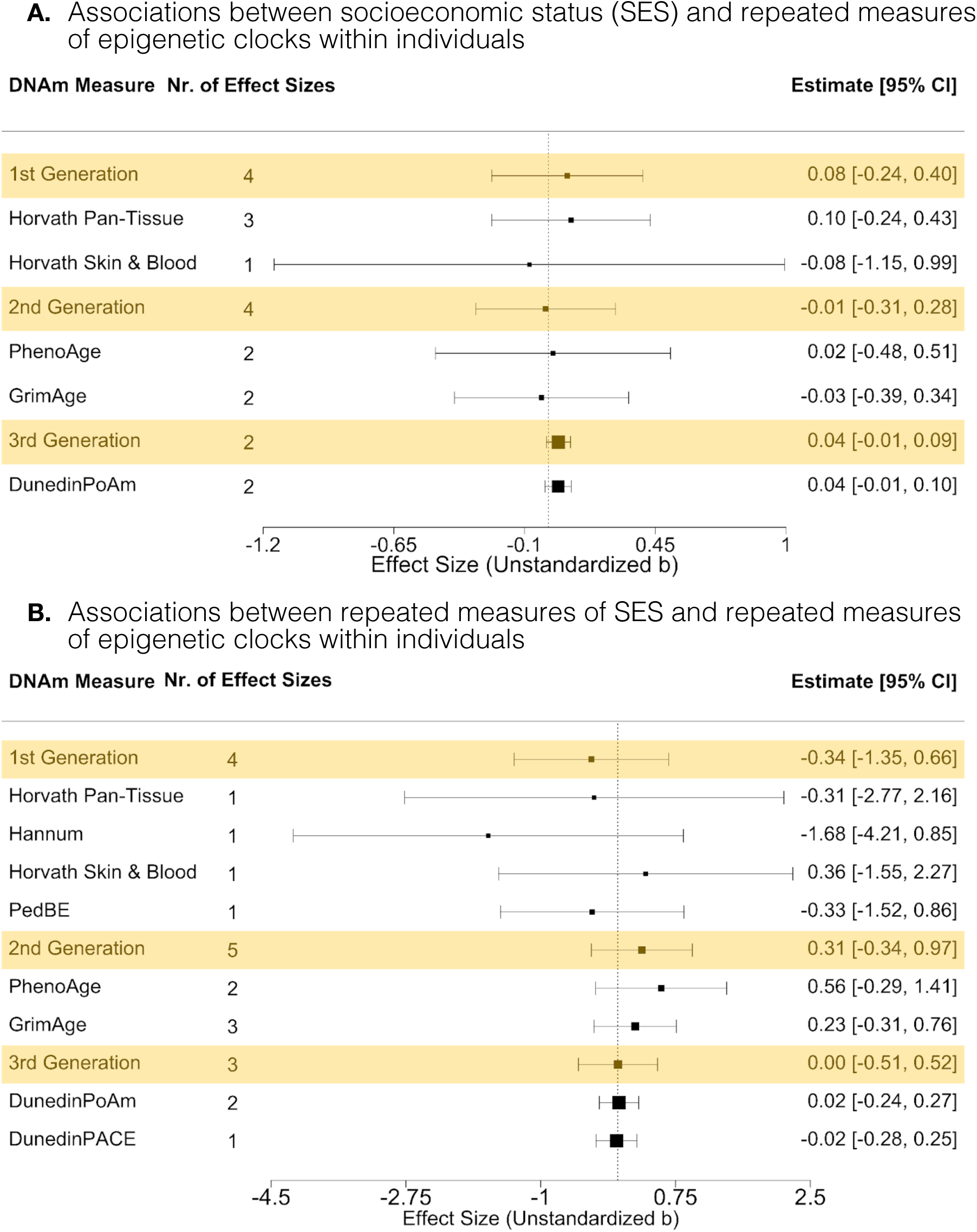
Associations between repeated measures of socioeconomic status (SES) and epigenetic clocks. Forest plot in section A depicts the meta-analytic associations between SES and repeated measures of epigenetic clocks within individuals. Forest plot in section B depicts the meta-analytic associations of repeated measures of SES and repeated measures of epigenetic clocks within individuals. Yellow panels indicate the meta-analyzed effect size estimates for first-, second-, and third-generation clocks, followed by individual clocks within each category. As effect sizes for change data are mainly reported as unstandardized regression coefficients, and SDs for change in age acceleration and SES are not reported in any of the studies, change results are meta-analyzed as unstandardized regression coefficients separately. Effect size estimates are bounded by 95% confidence intervals (CIs). * denotes significant difference from 0 at p < 0.05. The size of the boxes in the middle panel represents the precision of the effect size estimate (i.e., the smaller the CI, the more precise the estimate).

**Supplemental Figure 6.**
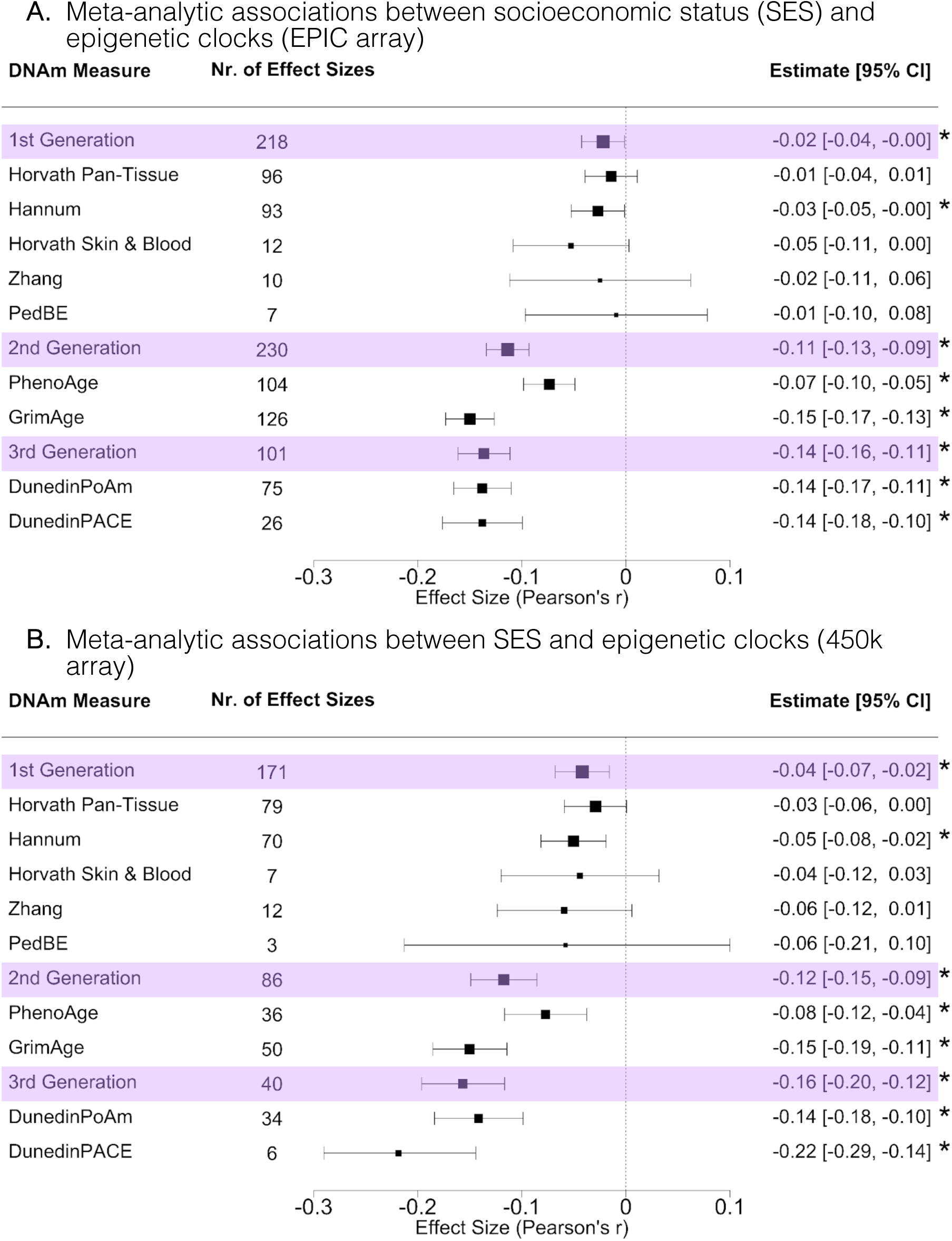
Meta-analytic associations between socioeconomic status (SES) and epigenetic clocks, separately for EPIC and 450k arrays. Forest plot depicts the meta-analytic associations of SES and epigenetic clocks for EPIC array (section A) and 450k array (section B). Purple panels indicate the meta-analyzed effect size estimates for first-, second-, and third-generation clocks, followed by individual clocks within each category. Analyses are conducted using Fisher-Z transformation and later transformed to Pearson’s r to ease interpretation. Effect size estimates are bounded by 95% confidence intervals (CIs). * denotes significant difference from 0 at p < 0.05. The size of the boxes in the middle panel represents the precision of the effect size estimate (i.e., the smaller the CI, the more precise the estimate).

**Supplemental Figure 7.**
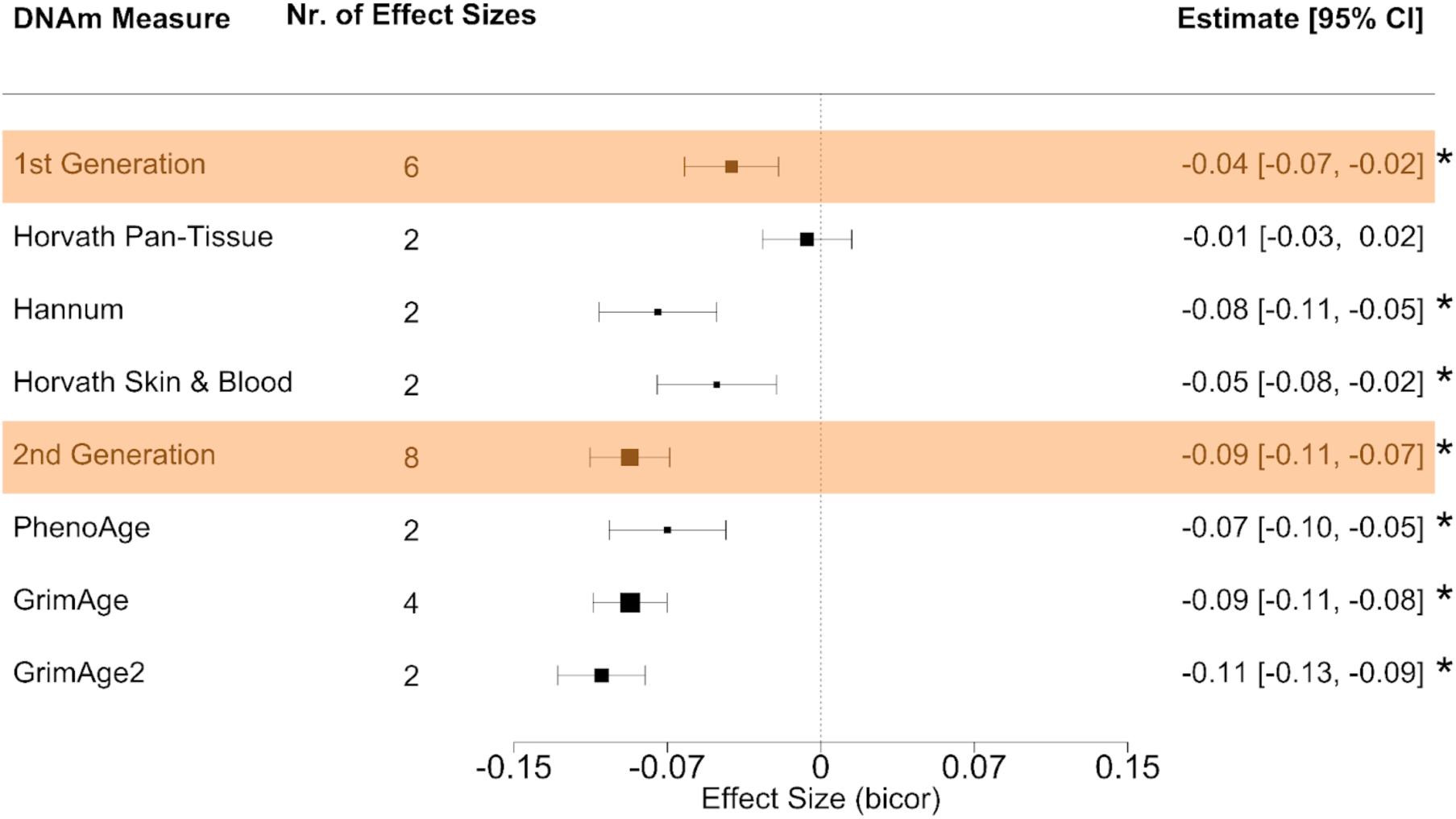
Meta-analytic associations between socioeconomic status (SES) and epigenetic clocks – biweight midcorrelation (bicor) estimates. Forest plot depicts the meta-analytic associations between SES and epigenetic clocks. As a reliable conversion from bicor to Pearson’s r could not be found, bicor effect sizes are analyzed separately. Orange panels indicate the meta-analyzed effect size estimates for first- and second-generations clocks, followed by individual clocks within each category. Effect size estimates are bounded by 95% confidence intervals (CIs). * denotes significant difference from 0 at p < 0.05. The size of the boxes in the middle panel represents the precision of the effect size estimate (i.e., the smaller the CI, the more precise the estimate).

